# Pronounced Regional Variation in Esketamine and Ketamine Prescribing to US Medicaid Patients

**DOI:** 10.1101/2022.04.23.22274206

**Authors:** Alexia G. Aguilar, Burke A. Beauregard, Christopher P. Conroy, Yashoda T. Khatiwoda, Shantia M. E. Horsford, Stephanie D. Nichols, Brian J. Piper

## Abstract

**Background:** Ketamine and esketamine are efficacious for treatment resistant depression. Unlike other antidepressants, ketamine and esketamine lack a therapeutic delay and do not increase risk for suicidal thoughts and behaviors in adolescents and young-adults. Esketamine gained FDA approval in March of 2019.

**Objective:** This cross-sectional study aimed to geographically characterize ketamine and esketamine prescribing to US Medicaid patients.

**Methods:** Ketamine and esketamine prescription rate and spending per state were obtained from the Medicaid State Drug Utilization Database. States outside of a 95% Confidence Interval were considered statistically significant.

**Results:** Between 2009-2020, ketamine prescribing rates peaked in 2013 followed by a general decline. For ketamine and esketamine in 2019, Montana (967/million enrollees) and Indiana (425) showed significantly higher prescription rates, respectively, relative to the national average. A total of 21 states prescribed neither ketamine nor esketamine in 2019. Since its approval, esketamine prescriptions have surpassed those of ketamine. There was a 121.3% increase in esketamine prescriptions from 2019 to 2020. North Dakota (1,423) and North Carolina (1,094) were significantly elevated for esketamine in 2020. Ten states prescribed neither ketamine nor esketamine in 2020. State Medicaid programs in 2020 spent 72.7 fold more for esketamine ($25.3 million) than on ketamine (0.3 million) prescriptions.

**Conclusion:** Despite the effectiveness of ketamine and esketamine for treatment resistant depression, their use among Medicaid patients was limited and variable in many areas of the US.

## Introduction

Suicide rates continued to increase in the United States marking the presence of a suicide epidemic. Suicide was the second leading cause of death for people aged 10-34 in 2019 [1]. Suicide ranked as the 10th leading cause of death with 4.8% of adults having thoughts of suicide and 0.6% (1,969,200) reporting having had attempted suicide in the past year [1]. Since the emergence of COVID-19, serious suicide attempts have increased in adolescent girls but not in boys [2]. The rate of serious suicide attempt ED visits increased by 50.6% in girls in 2021 compared with 2019 [2]. Among boys, the increase in serious suicide attempts was 3.7% [2]. Unfortunately, traditional antidepressants are often ineffective in preventing suicide linked to depression for three reasons. First, treatment resistant depression, or failure to respond to traditional antidepressants is present in up to two-thirds of patients. Second, a time lag of weeks to months delays the onset of therapeutic effects [3]. Third, traditional antidepressants have been found to counterintuitively increase the risk of suicidality in the first month of therapy in patients who are under 25. This effect was not found in adults and a protective effect was observed in geriatric adults, emphasizing age related differences in suicidality and response to these medications [4].

Ketamine, a well-known anesthetic agent, has gained attention as an antidepressant using a novel mechanism of action and is very useful for reducing suicidal ideation [5]. Its absence of therapeutic delay and effectiveness for treatment-resistant depression are well-suited for patients at risk of suicide [3]. By being effective in both major unipolar and bipolar depressive episodes, unlike other antidepressants, it is useful for depression even when the diagnostic picture is unclear. In March of 2019, the FDA issued a conditional approval of esketamine, a stereoisomer of ketamine, as nasal spray which is administered under direct supervision of a medical provider. It is approved for use in patients with treatment resistant depression or with depression and acute suicidality. A moderately well powered clinical trial (N = 63) demonstrated that esketamine (0.25 mg/kg iv) was non-inferior to ketamine (0.5 mg/kg iv) for treatment resistant depression [6]. The discovery of these new indications for ketamine is a landmark development as existing antidepressants have a black-box warning for young-adults for increasing suicide. Although Electroconvulsive Therapy (ECT) is a highly efficacious option for treatment resistant depression, it is generally recognized as underutilized [7], particularly among minorities [8-9].

The objective of this study was to examine patterns in ketamine and esketamine prescription rates throughout the United States among Medicaid patients. Determining geographic variability in prescription could be useful for correlating use of ketamine and esketamine to rates of suicide and beginning to examine corrective efforts on a population level. Given the recency of esketamine to the market and its current high visibility in the field, these patterns may be particularly informative.

## METHODS

### Procedures

Ketamine and esketamine prescription rates and costs were obtained from the Medicaid State Drug Utilization database [10]. Medicaid is a joint federal and state program that provides coverage for 75 million people, approximately 21% of the United States population. All states provide coverage for outpatient prescription drugs [10]. We evaluated the Medicaid State Drug Utilization database for total outpatient ketamine (2009 – 2020) and esketamine (2019 – 2020) prescriptions per state [10]. Prescription rates were reported per 1,000,000 Medicaid enrollees. Formulations were categorized by National Drug Codes (Supplemental Appendix A & B) by route of administration (nasal vs injection). Procedures were approved as exempt by the Geisinger and the University of New England IRBs.

### Data analysis

The following analyses were completed: 1) percent change in prescribing over time, 2) a 95% confidence interval (mean ± 1.96*SD) with states outside this range interpreted as significantly different from the mean, 3) the ratio of the highest to lowest (non-zero) prescribing rate, and 4) ratio of total Medicaid spending for esketamine relative to ketamine. We analyzed the data and constructed figures using SAS, JMP and GraphPad Prism.

## RESULTS

Figure 1 shows that prescribing rates for ketamine from 2009 to 2020 were elevated in 2013 with a 91.2% increase compared to 2012. This was followed by a gradual decrease from 2014 to 2018. With the approval of esketamine, the total (i.e. ketamine and esketamine) prescriptions tripled from 2018 (106/million enrollees) to 2020 (292/million). Examination of the quarterly prescriptions nationally revealed that ketamine was relatively stable except for a 31.2% reduction from the fourth quarter of 2019 (56.6) until the first quarter of 2020 (38.9). Esketamine increased 4.5-fold between the second quarter of 2019 (23.5) until the fourth quarter of 2020 (106.7, Supplemental Figure 1).

**Figure 1.**
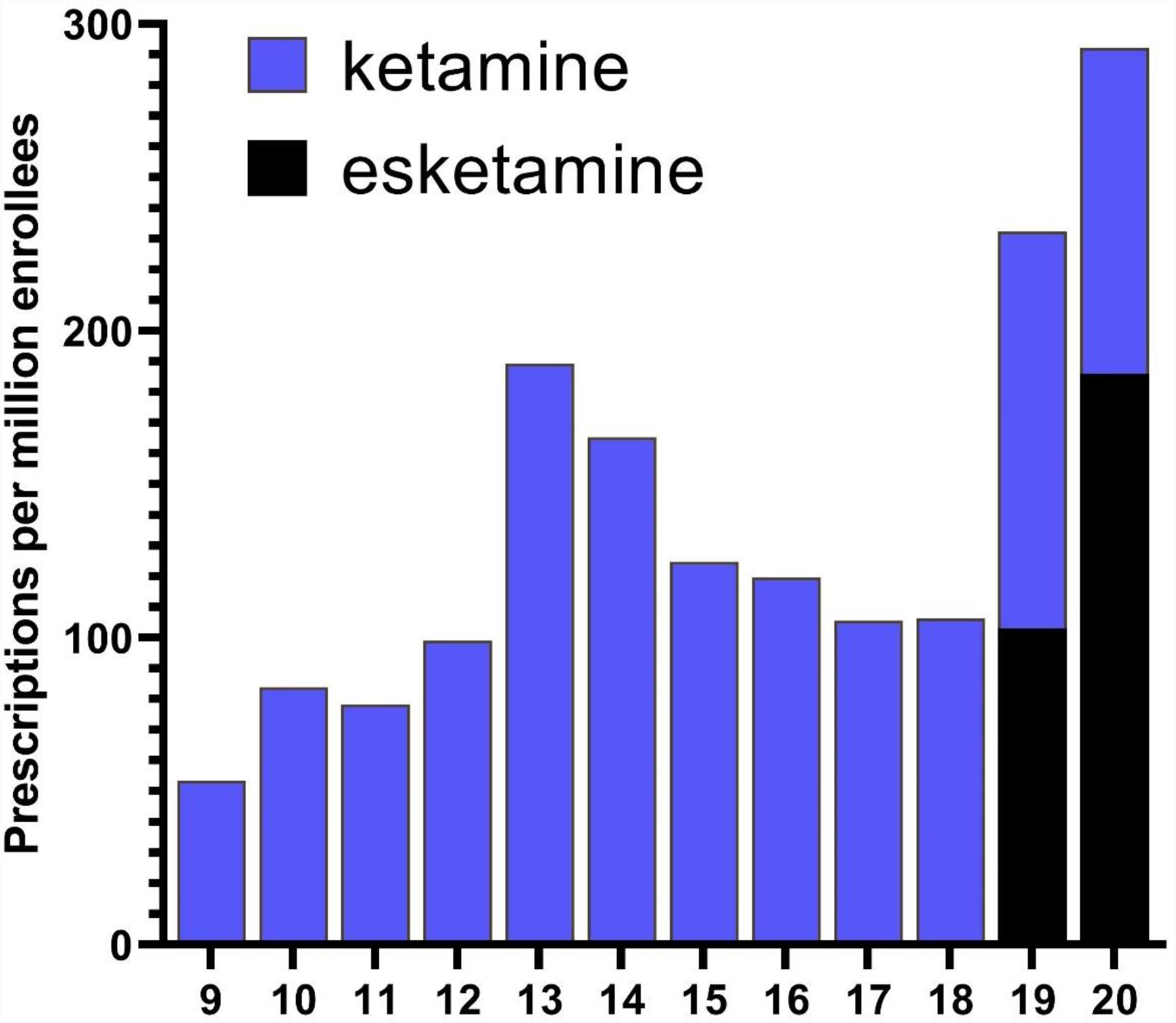
National prescription rate of ketamine and esketamine to Medicaid patients from 2009 to 2020.

There were 27,238 prescriptions nationally for ketamine and esketamine in 2019. Of these, 12,668 prescriptions (46.5%) were for esketamine nasal spray and 14,570 (53.5%) for ketamine injections. Pronounced geographical differences were also identified. For ketamine, Montana (967.3) showed a statistically significant (*p* < .05) higher rate of prescription relative to the national average (Figure 2A). There was a 116-fold higher rate of prescriptions in Montana relative to the lowest state (MA = 8.3). States in the southern US from New Mexico to South Carolina had no ketamine prescriptions (Supplemental Figure 1). For esketamine, Indiana (424.6) showed a significantly higher rate of prescription which was also 61-fold higher than the lowest state (Supplemental Figure 2). Twenty states (AK, AL, AR, DE, GA, HI, IA, ID, IL, KS, ME, MS, ND, NH, OK, RI, SC, VT, WI, WY) and Washington DC prescribed neither ketamine nor esketamine.

**Figure 2.**
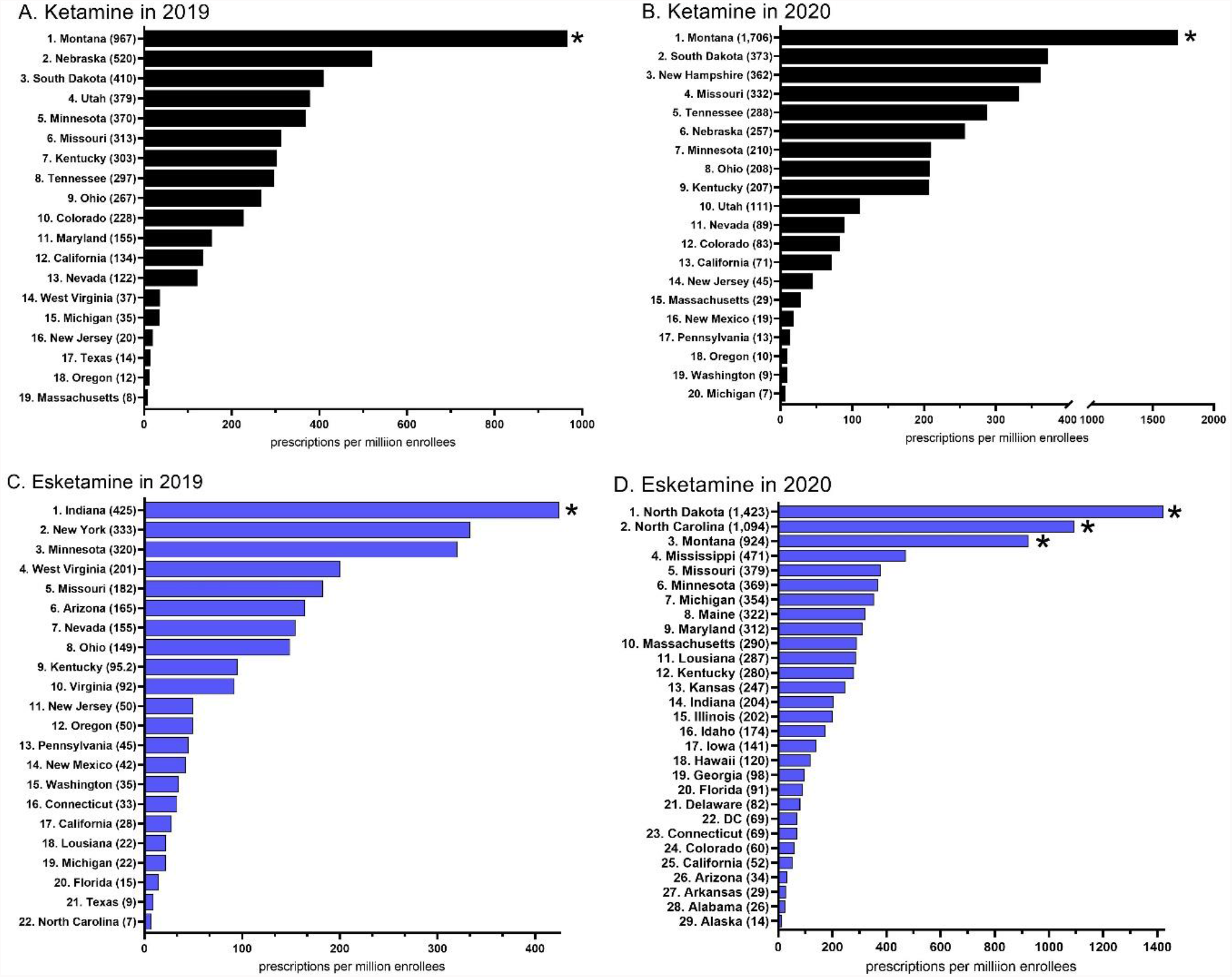
Ketamine (A, C) and esketamine (B, D) prescription rate per state to Medicaid patients. States not shown had a value of 0. * *p* < .05 vs the state average.

There were 40,765 prescriptions for ketamine and esketamine in 2020. Esketamine (28,037) was over double that of ketamine (12,728). Montana (1,705.6) was significantly higher than the national average and was 245-fold higher than the lowest state (Michigan = 7.0, Figure 2B) for ketamine. States that were located at or below the 35° latitude (Arizona proceeding east to South Carolina) had the lowest prescribing (Supplemental Figure 3). Three-states, North Dakota, North Carolina, and Montana were significantly elevated for esketamine in 2020. There was a 104-fold difference between the highest (North Dakota = 1,423.0) and lowest (Alaska = 13.7) states (Figure 2D, Supplemental Figure 4). Ten states (NY, OK, RI, SC, TX, VI, VT, WV, WI, & WY) prescribed neither ketamine nor esketamine.

The total spending by state Medicaid programs was 14.2 fold greater for esketamine ($5.1 million) than ketamine ($0.4 million) in 2019. This increased to 72.7 fold higher for esketamine ($25.3 million) than ketamine ($0.3 million) in 2020.

## DISCUSSION

There are two key findings from this report. First was the increase in esketamine relative to ketamine prescribing to Medicaid patients. Esketamine, in conjunction with an oral antidepressant, received FDA-approval for the treatment of adults with treatment-resistant depression on March 5, 2019. Esketamine prescriptions were less than half (44.2%) those of ketamine in 2019. The Sequenced Treatment Alternatives to Relieve Depression (STAR*D) study revealed that only one-third of depressed patients experienced a remission after receiving the first-line antidepressant [11]. Use of ketamine and esketamine when expressed relative to the number of Medicaid enrollees, and the ubiquity of depression, was modest. Second, the state-level differences in esketamine and ketamine prescription rates were pronounced. For example, one state, Indiana prescribed 25% of the national total esketamine in 2019. At the beginning of 2019, quarterly esketamine was prescribing was, as anticipated, quite limited, but as the year progressed, a subset of states quickly adapted to use of esketamine. This was perhaps due to increasing clinical evidence [3, 5, 6, 13] and wider distribution of trial results, and the rapid establishment of protocols and procedures that meet the FDA Risk Evaluation and Mitigation Strategy program requirements for these Schedule III substances. The pronounced differences in esketamine prescribing at a state-level could be attributed to marketing rather than long-standing preset preferences in patients and physicians since this was a new drug to the market.

The identification of the rapid therapeutic and anti-suicidality effects of ketamine is potentially one of the most important developments in psychopharmacology in the past two decades. However, we found that the use of esketamine and ketamine among Medicaid patients was both modest, relative to the prevalence of depression and treatment resistant depression, and extremely variable. This variability is also shown by the absence of ketamine/esketamine prescriptions in two-fifths of US states in an outpatient setting in 2019 and one-fifth of states in 2020. The role of ketamine and esketamine in the US as an antidepressant continues to evolve [6, 12-14].

Regardless of the underlying reasons, it is interesting that the ketamine and esketamine prescribing are nowhere near as high as one might expect from an antidepressant in a class on its own which overcomes the therapeutic lag of selective serotonin reuptake inhibitors and serotonin norepinephrine reuptake inhibitors. Among the top two-hundred most prescribed medications known for their antidepressant effects written for Medicaid patients in 2019 [10], fluoxetine was the most prescribed. The ratio of fluoxetine to ketamine prescription was nearly eight-hundred-fold different [10]. While it is not yet conclusive that a “miracle drug” has been found to treat depression, the rates of prescribing suggest peripheral challenges like the logistics of proper administration and/or social inertia on the use of this psychoactive and controlled substance that may be causing delays. The COVID-19 pandemic likely also contributed to the limited utilization of these agents. However, there have also been fifteen ketamine/esketamine manuscripts which were subsequently retracted (e.g. use of ketamine in the ER [18]) which may also be contributing to some ambiguity [Supplemental Appendix C]. There are also serious concerns with misuse and the development of tolerance [19].

The tremendous costs of esketamine/ketamine including monitoring post administration relative to more traditional treatments [20-22] may explain the hesitance among many state’s Medicaid programs to condone use of these agents. Esketamine prescriptions were double those of ketamine in 2020 while spending was seventy-three fold higher. Interesting, R-ketamine exerted a longer lasting antidepressant effect than S-ketamine in the mouse tail suspension test [23]. Similar findings of R-ketamine being more potent or producing longer duration effects than S-ketamine have also been observed in the forced swimming, learned helplessness, and social defeat stress models of depression [24-25]. A direct comparison revealed that three-fifths (62.1%) of patients had an antidepressant response to racemic ketamine relative to only two-fifths (43.7%) to esketamine at one-week although this difference was not significant [6]. The potential fiscal and therapeutic advantages of the racemate over only the S-enantiomer may warrant continued monitoring by those who oversee state Medicaid programs and others.

Some caveats and future directions are noteworthy. The Medicaid state drug utilization database does not provide information about patient medical history or demographics including race/ethnicity. Therefore, we can not quantify the portion of ketamine that was used for anesthetic purposes. There have been concerns that Blacks were half as likely to receive ECT as non-Hispanic whites [6, 7]. US hospitals in the South and West were less likely than those in the Northeast and Midwest to have ECT available [7]. Future research with electronic medical records will be necessary to evaluate whether these disparities are due to the same factors for ketamine and esketamine. However, it did not escape notice that the more diverse southern states also prescribed less ketamine (Texas, Arizona, Florida) and esketamine (Texas and Alabama). Follow-up investigations with Medicaid, Medicare, and private insurance should also be completed after the results of ketamine/esketamine and ECT comparisons [24-27] are completed.

In conclusion, this study found that low and extremely regionally disparate, rates of ketamine and esketamine prescribing to Medicaid patients in 2019 and 2020. It will likely be a fruitful endeavor to continue to monitor how use of these rapidly acting agents change in future years.

## Supporting information

Raw Medicaid Data

## Data Availability

All data produced are available online at:
https://www.medicaid.gov/medicaid/prescription-drugs/state-drug-utilization-data/index.html

## Acknowledgements

BJP was supported by the Health Resources Services Administration ((D34HP31025). Software used in this effort was provided by the NIEHS (T32-ES007060-31A1). Iris Johnston provided technical support.

---

**Supplemental Figure 1.**
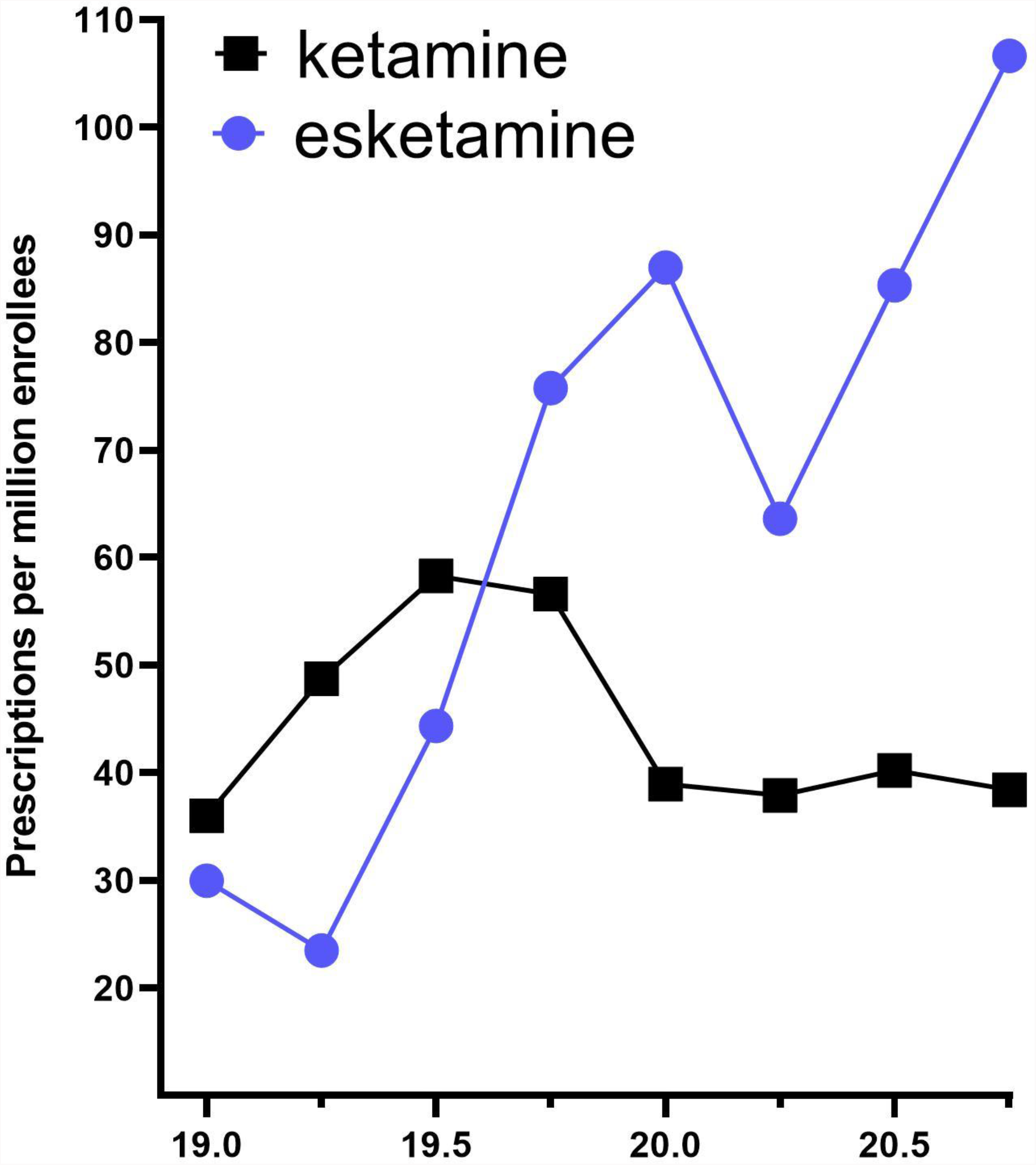
Quarterly prescriptions of ketamine and esketamine per million Medicaid enrollees in 2019 and 2020.

**Supplemental Figure 2.**
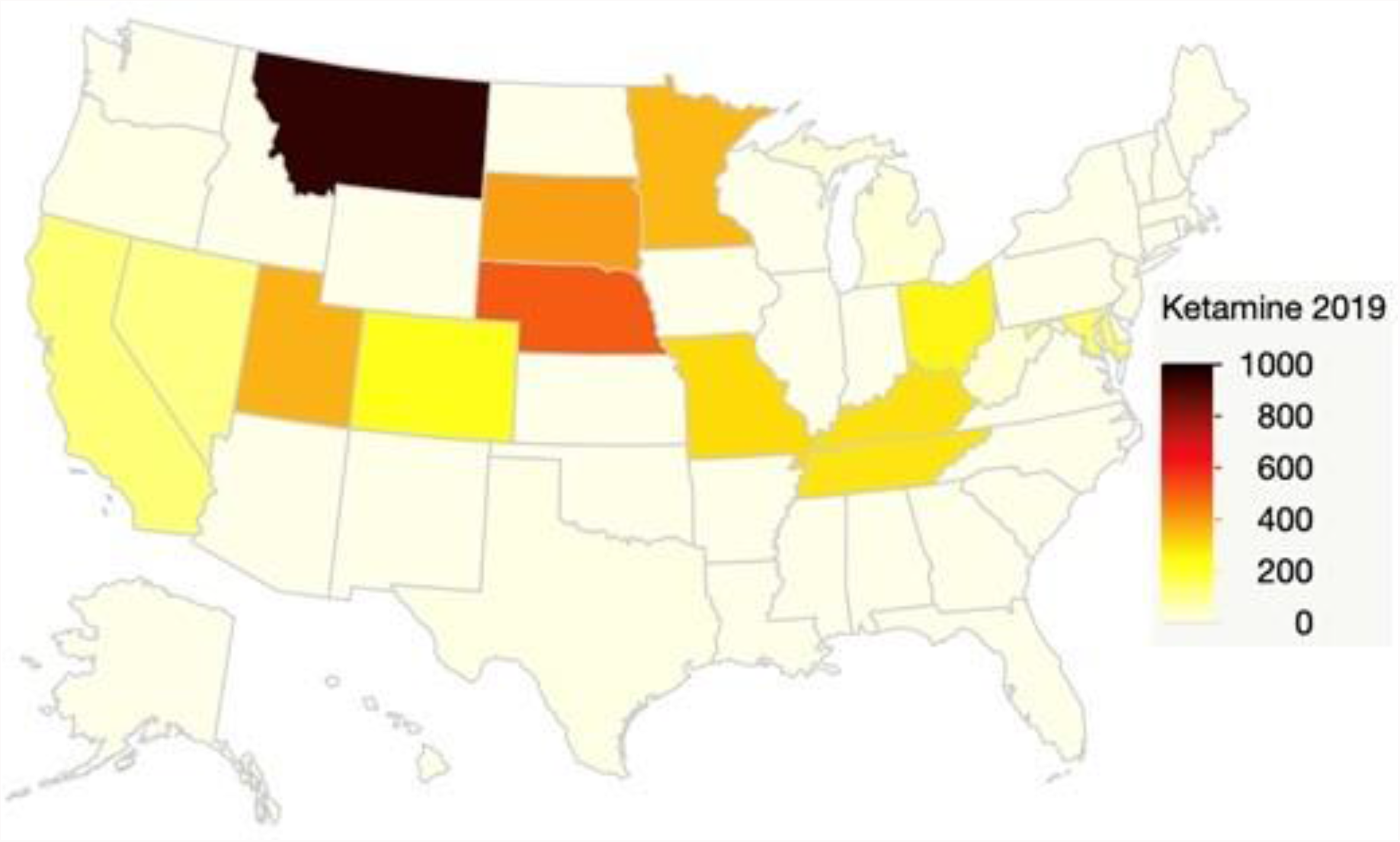
Heat map showing prescriptions of ketamine per million Medicaid enrollees in 2019.

**Supplemental Figure 3.**
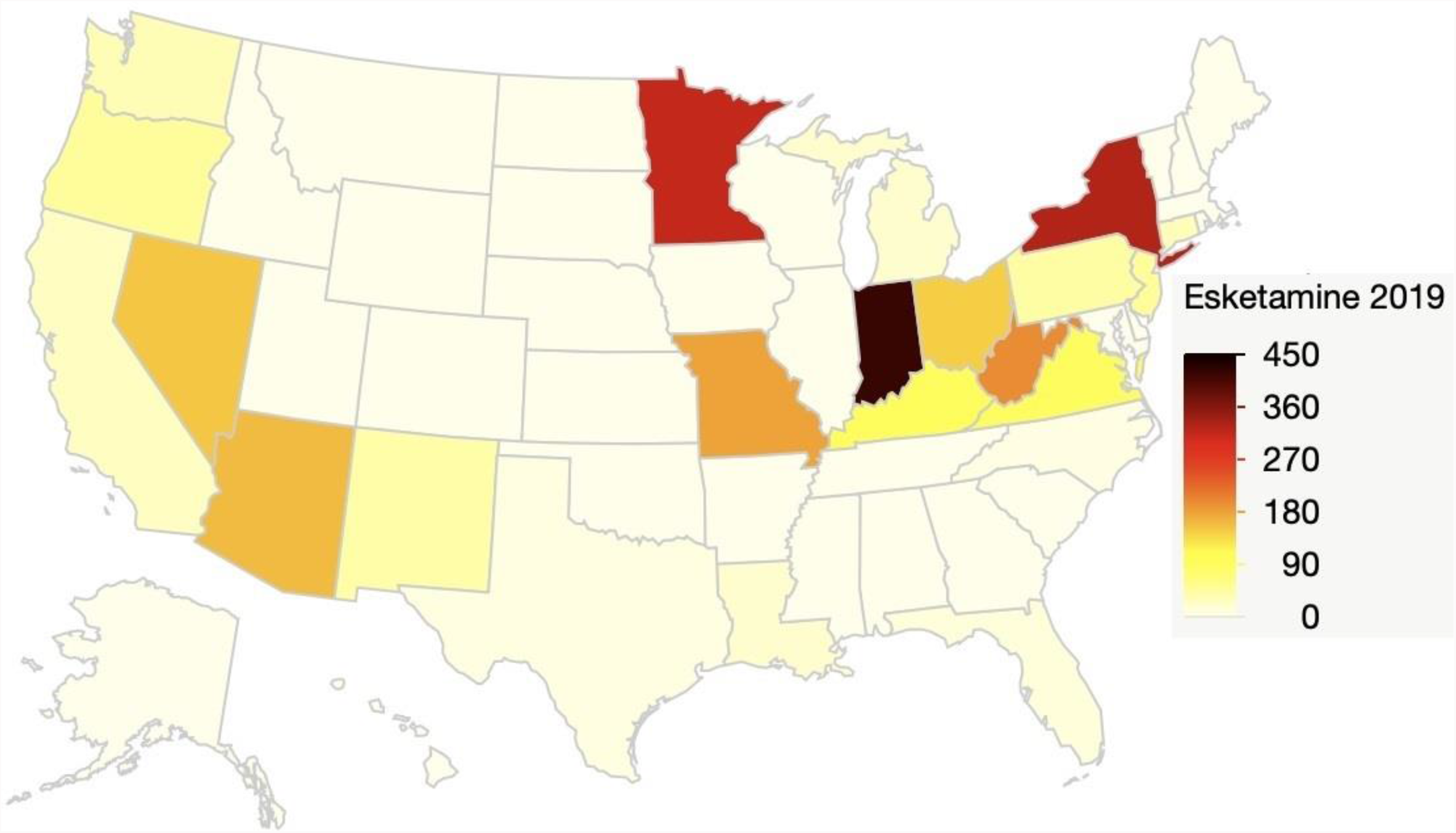
Heat map showing prescriptions of esketamine per million Medicaid enrollees in 2019.

**Supplemental Figure 4.**
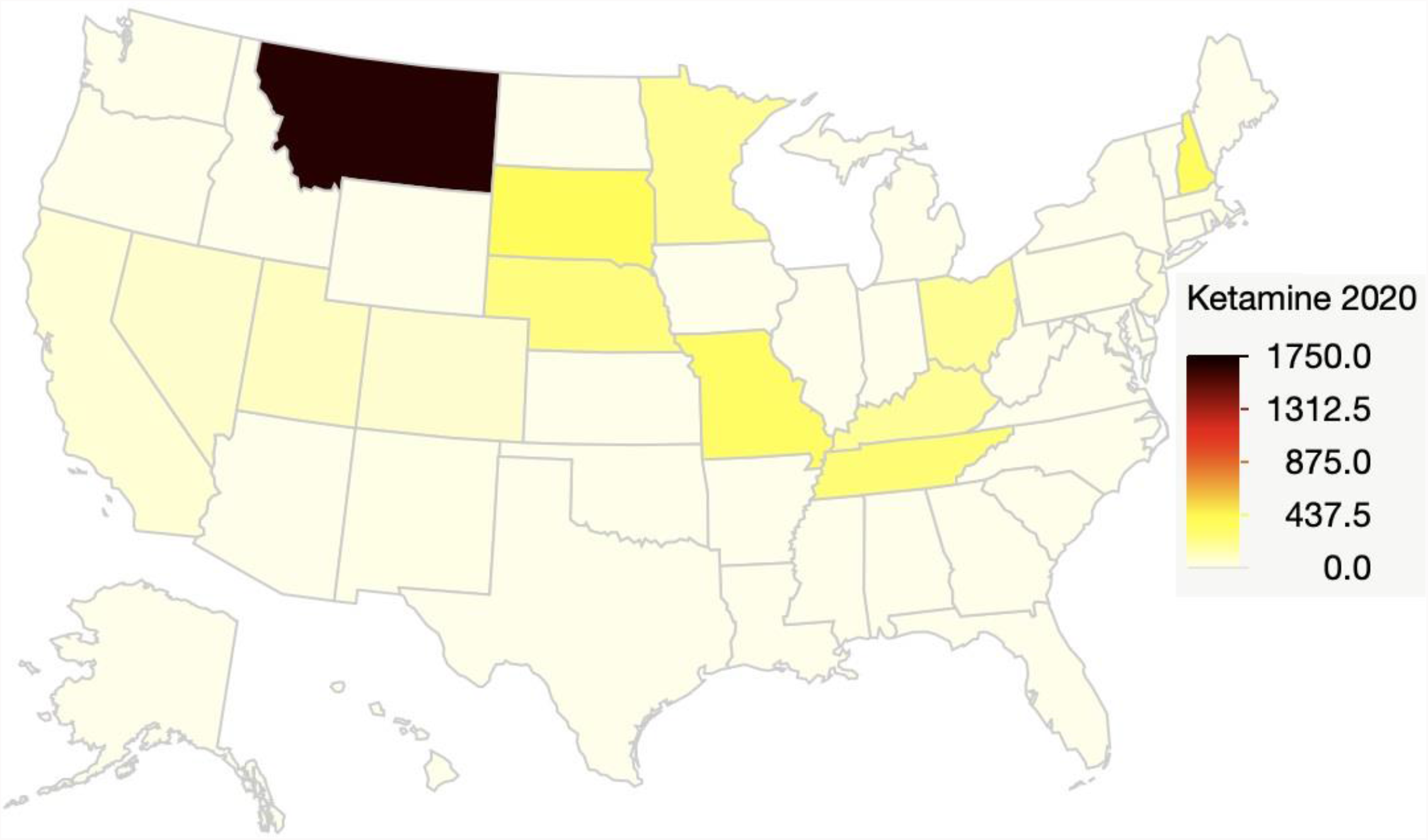
Heat map showing prescriptions of ketamine per million Medicaid enrollees in 2020.

**Supplemental Figure 5.**
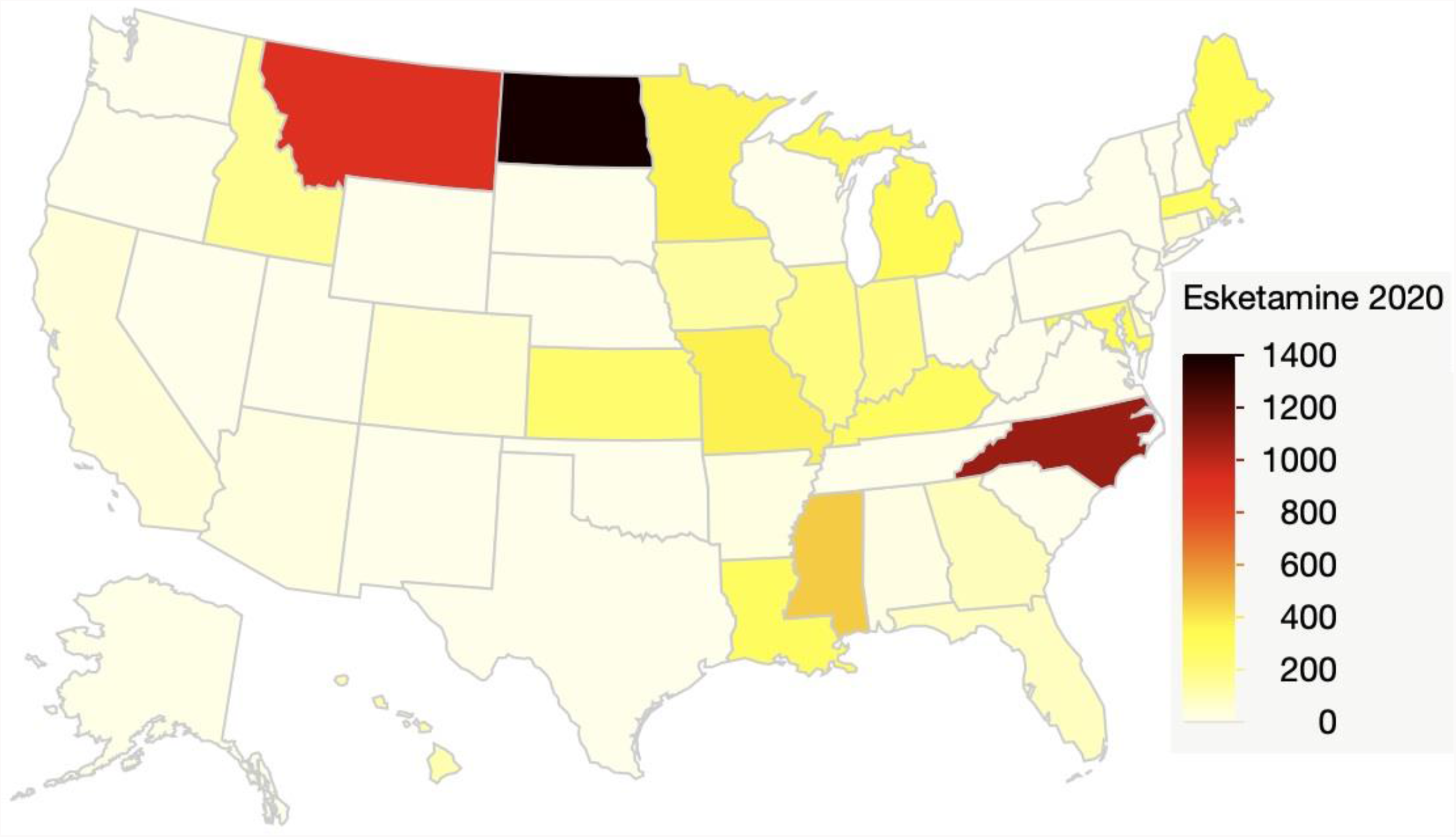
Heat map showing prescriptions of esketamine per million Medicaid enrollees in 2020.

### Supplementary Appendix A

Ketamine product name, National Drug Codes (NDC), and number of Medicaid prescriptions in 2019.

| <u>Product Name</u> | <u>NDC</u> | <u>Number of prescriptions</u> |
| --- | --- | --- |
| Ketamine 5 | 67457000110 | 2,139 |
|  | 409205105 | 851 |
|  | 143950810 | 659 |
|  | 409205310 | 468 |
|  | 143950801 | 374 |
|  | 67457000100 | 362 |
|  | 143950910 | 137 |
|  | 143950901 | 97 |
| Ketamine H | 67457000110 | 1,099 |
|  | 409205105 | 456 |
|  | 143950810 | 375 |
|  | 409205310 | 306 |
|  | 67457018100 | 293 |
|  | 67457018120 | 235 |
|  | 67457000100 | 67 |
|  | 67457010800 | 54 |
|  | 143950801 | 36 |
|  | 143950910 | 34 |
|  | 143950901 | 14 |
|  | 42023013810 | 14 |
|  | 42023013710 | 0 |
|  | 42023013910 | 0 |
|  | 67457010810 | 0 |
| Ketamine 50 | 42023011410 | 1,065 |
|  | 42023011510 | 801 |
| Ketalar 20 | 42023011310 | 1530 |
| Ketalar C- | 42023011310 | 898 |
|  | 42023011410 | 408 |
|  | 42023011510 | 223 |
| Ketalar | 42023011310 | 204 |
|  | 42023011510 | 180 |
|  | 42023011410 | 56 |
| Ketamine 2 | 67457018120 | 740 |
|  | 67457018100 | 222 |
| Ketamine 1 | 67457010810 | 81 |
|  | 409205105 | 80 |
|  | 67457010800 | 12 |

### Supplemental Appendix B

Eskatamine product name and National Drug Code (NDC), and Medicaid prescriptions in 2019.

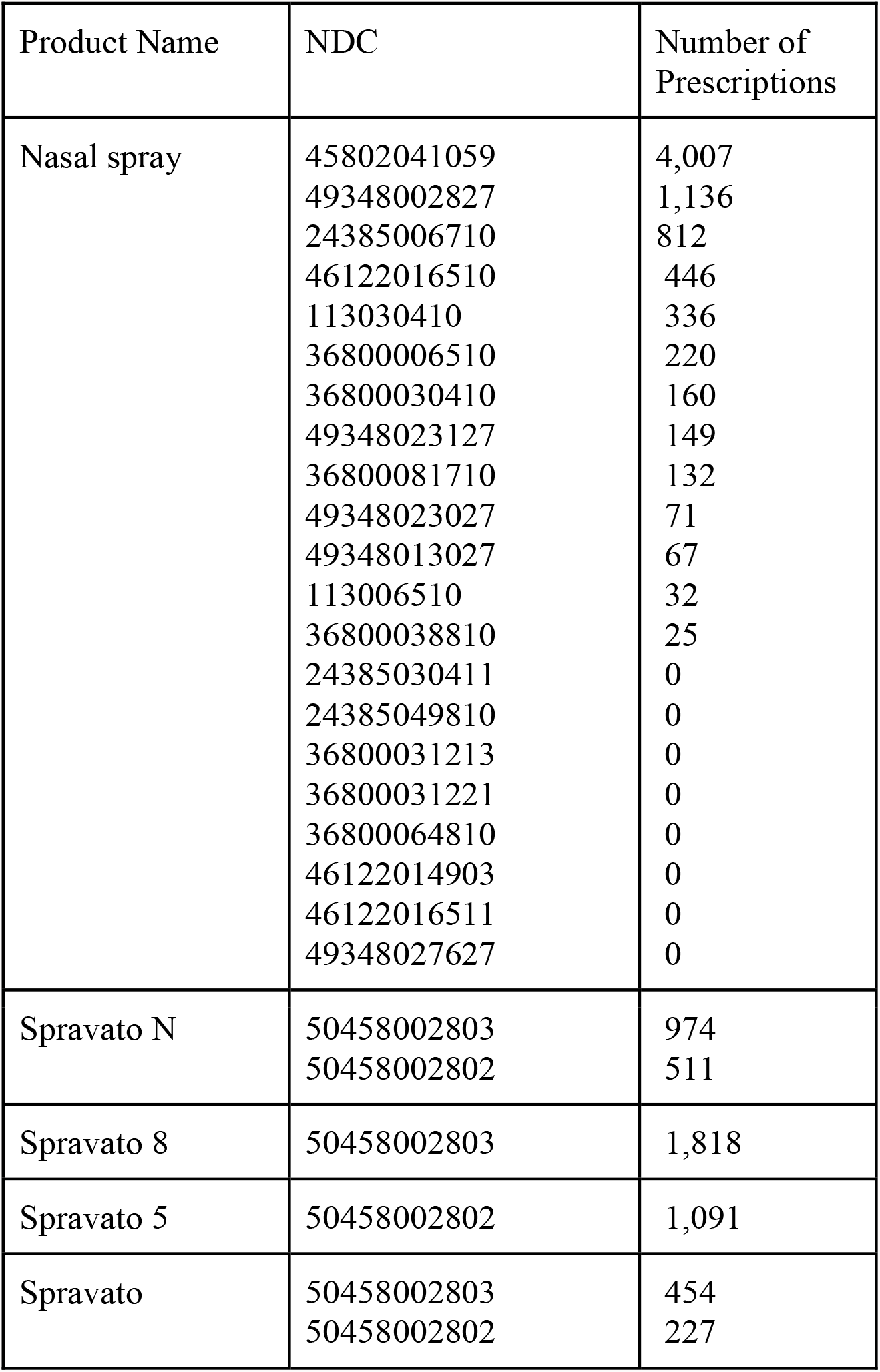

**Supplemental Appendix B.** Eskatamine product name and National Drug Code (NDC), and Medicaid prescriptions in 2019.
| Product Name | NDC | Number of Prescriptions |
| --- | --- | --- |
| Nasal spray | 45802041059 | 4,007 |
|  | 49348002827 | 1,136 |
|  | 24385006710 | 812 |
|  | 46122016510 | 446 |
|  | 113030410 | 336 |
|  | 36800006510 | 220 |
|  | 36800030410 | 160 |
|  | 49348023127 | 149 |
|  | 36800081710 | 132 |
|  | 49348023027 | 71 |
|  | 49348013027 | 67 |
|  | 113006510 | 32 |
|  | 36800038810 | 25 |
|  | 24385030411 | 0 |
|  | 24385049810 | 0 |
|  | 36800031213 | 0 |
|  | 36800031221 | 0 |
|  | 36800064810 | 0 |
|  | 46122014903 | 0 |
|  | 46122016511 | 0 |
|  | 49348027627 | 0 |
| Spravato N | 50458002803 | 974 |
|  | 50458002802 | 511 |
| Spravato 8 | 50458002803 | 1,818 |
| Spravato 5 | 50458002802 | 1,091 |
| Spravato | 50458002803 | 454 |
|  | 50458002802 | 227 |

### Supplementary Appendix C

Retracted ketamine articles reported to the RetractionWatch database. Two papers that were subsequently corrected are also listed. Rows highlighted in green are especially relevant to depression or bipolar treatment. As of 4/24/2022, there were no entries for esketamine.

| <u>Title</u> | <u>Journal</u> | <u>Outcome</u> |
| --- | --- | --- |
| Nardostachys jatamansi targets BDNF-TrkB to alleviate ketamine-induced schizophrenia-like symptoms in rats | *Neuropsychobiology* | Retracted Sept 13, 2021 |
| The sustained antidepressant effects of ketamine are independent of the lateral habenula | *Journal of Neuroscience* | Retracted May 5, 2021 |
| Ketamine reduces LPS-induced HMGB1 through HO-1 and Nrf2/p38 MAPK | *Journal of Surgical Research* | Retracted November, 2020 |
| Negative trial of scopolamine in major depressive disorder does not demonstrate neurophysiological changes seen with the antidepressant response of ketamine | *Biological Psychiatry* | Retracted May 1, 2018 |
| Efficacy of ketamine in bipolar depression: Systematic review and meta-analysis | *Journal of Psychiatric Practice* | Retracted July, 2018 |
| The influence of ketamine on the locus coeruleus and the noradrenergic system- a randomized fMRI study | *Clinical Neurophysiology* | Retracted Sept 4, 2018 |
| A preliminary naturalistic study of low-dose ketamine for depression and suicide ideation in the emergency department | *International Journal of Neuropsychopharmacology* | Retracted July 1, 2017 |
| Antidepressant potential of (R)-ketamine in rodent models: Comparison with (S)-ketamine. | *Journal of Pharmacology and Experimental Therapeutics* | Correction April 2017 |
| R-ketamine: a rapid-onset and sustained antidepressant without psychotomimetic side effects | *Translational Psychiatry* | Correction August 20, 2020 |
| Postoperative analgesedation with S(+) ketamine decreased the incidences of postanesthetic shivering and nausea and vomiting after cardiac surgery | *Medical Science Monitor* | Retracted Dec 1, 2008 |
| Perioperative ketamine for acute postoperative pain | *Cochrane Database of Systematic Reviews* | Retracted (withdrawn) July 2, 2015 |
| A randomized, double-blind, placebo controlled study assessing the anti-inflammatory effects of ketamine in cardiac surgical patients | *Journal of Cardiothoracic and Vascular Anesthesia* | Retracted Oct, 2014 |
| Efficacy of lignocaine plus ketamine at different doses in the prevention of pain due to propofol injection | *Clinical Drug Investigation* | Retracted May, 2014 |
| Sevoflurane-emergence agitation: Effect of supplementary low-dose oral ketamine premedication in preschool children undergoing dental surgery | *European Journal of Anaesthesiology* | Retracted April, 2011 |
| Assessment of recovery, dreaming, hemodynamics, and satisfaction in post cardiac surgery patients receiving supplementary propofol sedation with S(+)- ketamine | *Minerva Anestesiologica* | Retracted May, 2011 |

